# Estimating the generation time for influenza transmission using household data in the United States

**DOI:** 10.1101/2024.08.17.24312064

**Authors:** Louis Yat Hin Chan, Sinead E. Morris, Melissa S. Stockwell, Natalie M. Bowman, Edwin Asturias, Suchitra Rao, Karen Lutrick, Katherine D. Ellingson, Huong Q. Nguyen, Yvonne Maldonado, Son H. McLaren, Ellen Sano, Jessica E. Biddle, Sarah E. Smith-Jeffcoat, Matthew Biggerstaff, Melissa A. Rolfes, H. Keipp Talbot, Carlos G. Grijalva, Rebecca K. Borchering, Alexandra M. Mellis, RVTN-Sentinel Study Group

**Affiliations:** Centers for Disease Control and Prevention; Columbia University Irving Medical Center; University of North Carolina at Chapel Hill; University of Colorado School of Medicine and Children’s Hospital Colorado; University of Arizona; Marshfield Clinic Research Institute; Stanford University; Vanderbilt University Medical Center; Goldbelt Professional Services

**Keywords:** Generation interval, Serial interval, Incubation period, Pre-symptomatic transmission, Household transmission, Respiratory diseases

## Abstract

The generation time, representing the interval between infections in primary and secondary cases, is essential for understanding and predicting the transmission dynamics of seasonal influenza, including the real-time effective reproduction number (Rt). However, comprehensive generation time estimates for seasonal influenza, especially post the 2009 influenza pandemic, are lacking.

We estimated the generation time utilizing data from a 7-site case-ascertained household study in the United States over two influenza seasons, 2021/2022 and 2022/2023. More than 200 individuals who tested positive for influenza and their household contacts were enrolled within 7 days of the first illness in the household. All participants were prospectively followed for 10 days completing daily symptom diaries and collecting nasal swabs, which were tested for influenza via RT-PCR. We analyzed these data by modifying a previously published Bayesian data augmentation approach that imputes infection times of cases to obtain both intrinsic (assuming no susceptible depletion) and realized (observed within household) generation times. We assessed the robustness of the generation time estimate by varying the incubation period, and generated estimates of the proportion of transmission before symptomatic onset, infectious period, and latent period.

We estimated a mean intrinsic generation time of 3.2 (95% credible interval, CrI: 2.9-3.6) days, with a realized household generation time of 2.8 (95% CrI: 2.7-3.0) days. The generation time exhibited limited sensitivity to incubation period variation. Estimates of the proportion of transmission that occurred before symptom onset, the infectious period, and the latent period were sensitive to variation in incubation periods.

Our study contributes to the ongoing efforts to refine estimates of the generation time for influenza. Our estimates, derived from recent data following the COVID-19 pandemic, are consistent with previous pre-pandemic estimates, and will be incorporated into real-time Rt estimation efforts.

## Introduction

The generation time, a crucial parameter in understanding the dynamics of infectious diseases, is defined as the time interval between infections in primary and secondary cases. In the context of seasonal influenza, estimation of the generation time becomes increasingly important for predicting the trajectory of outbreaks and informed public health decision-making during an influenza season. This interval represents the time between typical influenza infections, reflecting when most transmission is likely to happen.

Estimating the generation time is challenging because few investigations can accurately detect the exact time of infection. The generation time is often inferred from the serial interval, defined as the time between symptom onsets of primary and secondary cases (Svensson 2007), due to the practicality of observing symptom onsets rather than infections. However, this alternative measure may not always approximate the generation time due to its dependence on the incubation period, defined as the duration from infection to symptom onset, and the possibility of asymptomatic infections.

Accurate estimation of the generation time is important for predicting the real-time effective reproduction number (Rt), a metric used to describe transmission intensities through time (Gostic, et al. 2020). During the 2023/2024 influenza season, the Centers for Disease Control and Prevention (CDC) estimated the current epidemic growth status for influenza infections in the U.S. as either growing or declining based on the Rt (Centers for Disease Control and Prevention 2024) (Centers for Disease Control and Prevention 2024). For this estimate, the generation time was approximated with a serial interval from a study by Cowling et al. (Cowling, et al. 2009) that utilized data collected in Hong Kong in 2007, prior to the 2009 H1N1 influenza pandemic.

To improve our understanding of seasonal influenza outbreaks, there is a need for more contemporary generation time estimates, especially following the COVID-19 pandemic. This analysis provides updated generation time estimates derived from an influenza household transmission study (Rolfes, et al. 2023) conducted during the 2021/2022 and 2022/2023 influenza seasons in the U.S. We employ a model using a published Bayesian data augmentation approach (Hart, Abbott, et al. 2022) (Hart, Maini and Thompson 2021) (Hart, Miller, et al. 2022) to impute missing event times, including infections and symptom onsets of cases, and estimate generation times. We estimate both the intrinsic generation time, which assumes no susceptible depletion, as well as the realized household generation time observed within the household setting. We also estimate the serial interval. We derived estimates across the two seasons, virus types (influenza A and B), and household sizes to understand potential differences and robustness to model assumptions. The insights gained from these sensitivity analyses contribute to our understanding of the reliability of our estimates across different data stratifications and assumptions, providing evidence that the generation time has remained substantially unchanged over the last decade or two.

We also estimate other transmission parameters, including the proportion of transmission before symptomatic onset, the infectious period, and the latent period. This helps in assessing pre- and post-symptomatic transmission, thereby providing insights to inform effective disease control strategies. These insights are crucial for preventing pre-symptomatic transmission through interventions such as isolation.

## Material and methods

### Household data

Participants included in this analysis were enrolled in a 7-site case-ascertained household study, the Respiratory Virus Transmission Network – Sentinel (RVTN-S), conducted in the U.S. over two consecutive influenza seasons: 2021/2022 and 2022/2023 (Rolfes, et al. 2023). After informed consent was obtained, the study enrolled individuals identified with influenza infections via polymerase chain reaction (PCR) testing and their household contacts within 7 days of the initial illness onset within the household.

Households were only enrolled if the index case who first presented for clinical testing was the first symptomatic or positive person in the household, with no other members of the household symptomatic on the first day of index case symptoms. Participants, including both index cases and household contacts, were then prospectively followed for 10 days, during which they completed daily symptom diaries and collected daily nasal swabs, which were tested for influenza via RT-PCR.

The dataset encompasses detailed information regarding symptoms and viral testing, including four main variables used in the model: whether individuals tested positive for influenza, their symptomatic status, dates of positive test results, and dates of symptom onset. Using the test positivity and symptomatic status, we stratified individuals into three types: symptomatic infected, asymptomatic infected, and uninfected. Both the dates of positive test results and symptom onset were used as upper bounds for the date of infection for each individual.

In the primary analysis, we excluded households with multiple co-primary cases, i.e., more than one individual exhibiting the same date of the earliest symptom onset concurrently. To assess robustness, we also performed a separate stratified analysis that included households both with and without multiple co-primary cases.

### Estimating the generation time

We employed a Susceptible-Exposed-Infectious-Recovered (SEIR) model with Bayesian data augmentation, originally developed by Hart et al. (Hart, Maini and Thompson 2021) for analyzing COVID-19 contact tracing data. The model was also used in two subsequent studies of household data in the United Kingdom (Hart, Abbott, et al. 2022) (Hart, Miller, et al. 2022). The SEIR model, referred to as the mechanistic model, which includes compartments for asymptomatic, pre-symptomatic and symptomatic infectious stages (Hart, Maini and Thompson 2021). Each stage may have different relative infectiousness, or transmission rates. Upon infection and entry into the non-infectious exposed stage, individuals may progress to become infectious through one of two pathways: either by remaining asymptomatic or by developing symptoms following a pre-symptomatic stage. Consequently, transmissions may occur before symptom onset, depending on the length of the incubation period.

We estimated both intrinsic and realized generation times by integrating data augmentation Markov Chain Monte Carlo (MCMC) techniques (Hart, Abbott, et al. 2022), to impute infection times and symptom onset of cases. The intrinsic generation time assumes no depletion of susceptible individuals, providing an estimate of the time it takes for an infected individual to infect others in the community with an unlimited supply of susceptible individuals. The realized household generation time reflects the actual time interval observed within households, restricted by the depletion of susceptible individuals over time. Susceptible depletion refers to the gradual reduction in the number of individuals within a population that have not yet been infected with a virus. For example, within the SEIR framework, members may become infected, develop immunity, and subsequently be removed from the susceptible pool. Considering this distinction allows for a more thorough understanding of influenza transmission dynamics, capturing both theoretical and observed aspects of transmission.

We also estimated several other crucial transmission parameters, including the proportion of transmission before symptomatic onset, the ratio of pre-symptomatic to symptomatic transmission rates (i.e., relative infectiousness of symptomatic infected individuals before symptom onset compared to after), as well as the latent period, the pre-symptomatic infectious period, and the symptomatic infectious period, all under the SEIR framework.

In adapting the model for our influenza study, we used estimates for the incubation period of influenza A from a systematic review by Lessler et al. (Lessler, et al. 2009). In sensitivity analyses, we explored variations derived from parallel estimates for influenza B (Lessler, et al. 2009) and for influenza A(H1N1)pdm09 (Tuite, et al. 2010).

Regarding the relative infectiousness of asymptomatic infected individuals compared with symptomatic infected individuals, we assumed a value of 0.57 (i.e., asymptomatic infected individuals were 43% less infectious than those symptomatic infected) based on the mean estimate from a recent study (Tsang, et al. 2023), and we also conducted sensitivity analyses using values of 0.11 and 1.54 based on the corresponding 95% credible interval (CrI).

To compare posterior distributions of estimates, we calculated the overlapping index, a measure of distribution similarities (Pastore 2018) (Pastore and Calcagnì 2019). A value close to 1 indicates high similarity, implying no substantial differences, while a value close to 0 indicates low similarity, implying substantial differences. We compared the estimates of generation time across multiple data stratifications and sensitivity analyses to the primary results excluding households with multiple co-primary cases.

The model was implemented in R (version 4.3.1) with 1,000,000 Markov chain Monte Carlo iterations, discarding the initial 20% as burn-in and obtaining posterior distributions by thinning every 100 iterations. The code for the model is available at https://github.com/CDCgov/influenza-generation_time-us.

### Ethics statement

This study was reviewed and approved by the IRB at Vanderbilt University Medical Center (see 45 C.F.R. part 46.114; 21 C.F.R. part 56.114).

## Results

### The household data

During the data cleaning process, we excluded 93 individuals who did not have at least two valid PCR tests and 2 individuals who were the only household members. In the primary analysis, we further excluded 23 individuals from 6 households that had co-primary cases. The final cleaned dataset, covering both seasons, comprised 820 individuals from 246 households (Table 1).

**Table 1.** Characteristics of household data.

| Data stratifications | Number of individuals (households) | Symptomatic infected % | Asymptomatic infected % | Uninfected % |
| --- | --- | --- | --- | --- |
| All data excluding households with multiple co-primary cases (primary analysis) | 820 (246) | 59.4% (487/820) | 7.2% (59/820) | 33.4% (274/820) |
| Season 2021/2022 | 308 (90) | 59.4% (183/308) | 7.5% (23/308) | 33.1% (102/308) |
| Season 2022/2023 | 512 (156) | 59.4% (304/512) | 7.0% (36/512) | 33.6% (172/512) |
| Influenza A | 683 (209) | 61.1% (417/683) | 7.5% (51/683) | 31.5% (215/683) |
| Influenza B | 137 (37) | 51.1% (70/137) | 5.8% (8/137) | 43.1% (59/137) |
| Household size of 2 or 3 | 393 (152) | 62.6% (246/393) | 5.3% (21/393) | 32.1% (126/393) |
| Household size of 4 or greater | 427 (94) | 56.4% (241/427) | 8.9% (38/427) | 34.7% (148/427) |
| All data including households with multiple co-primary cases | 843 (252) | 60.3% (508/843) | 7.0% (59/843) | 32.7% (276/843) |

As shown in Table 1, more households were enrolled in the 2022/2023 season. In both seasons, influenza A viruses predominantly circulated. In the 2021/2022 season, influenza A(H3N2) virus was identified in 78% of individuals, and influenza A(H1N1) virus was identified in 1% of individuals. In the 2022/2023 season, the percentages changed to 64% and 6%, respectively. Since households consisting of 3 or 4 members were the majority, we stratified the data into two groups: those with 2 or 3 members, and those with 4 or greater, to ensure comparability in quantity.

### Consistent estimates of the generation time across data stratifications and parameter assumptions

In the primary analysis using all data excluding households with multiple co-primary cases from both seasons, we estimated a mean intrinsic generation time of 3.2 (95% credible interval, CrI: 2.9-3.6) days (Figure 1A, 1B and Table 2). The corresponding mean (intrinsic) serial interval was 3.2 (95% CrI: 2.8-3.5) days, with a standard deviation (SD) of 2.2 (95% CrI: 1.8-2.6) days. The mean realized household generation time was 2.8 (95% CrI: 2.7-3.0) days, nearly half a day shorter than the mean intrinsic generation time.

**Figure 1.**
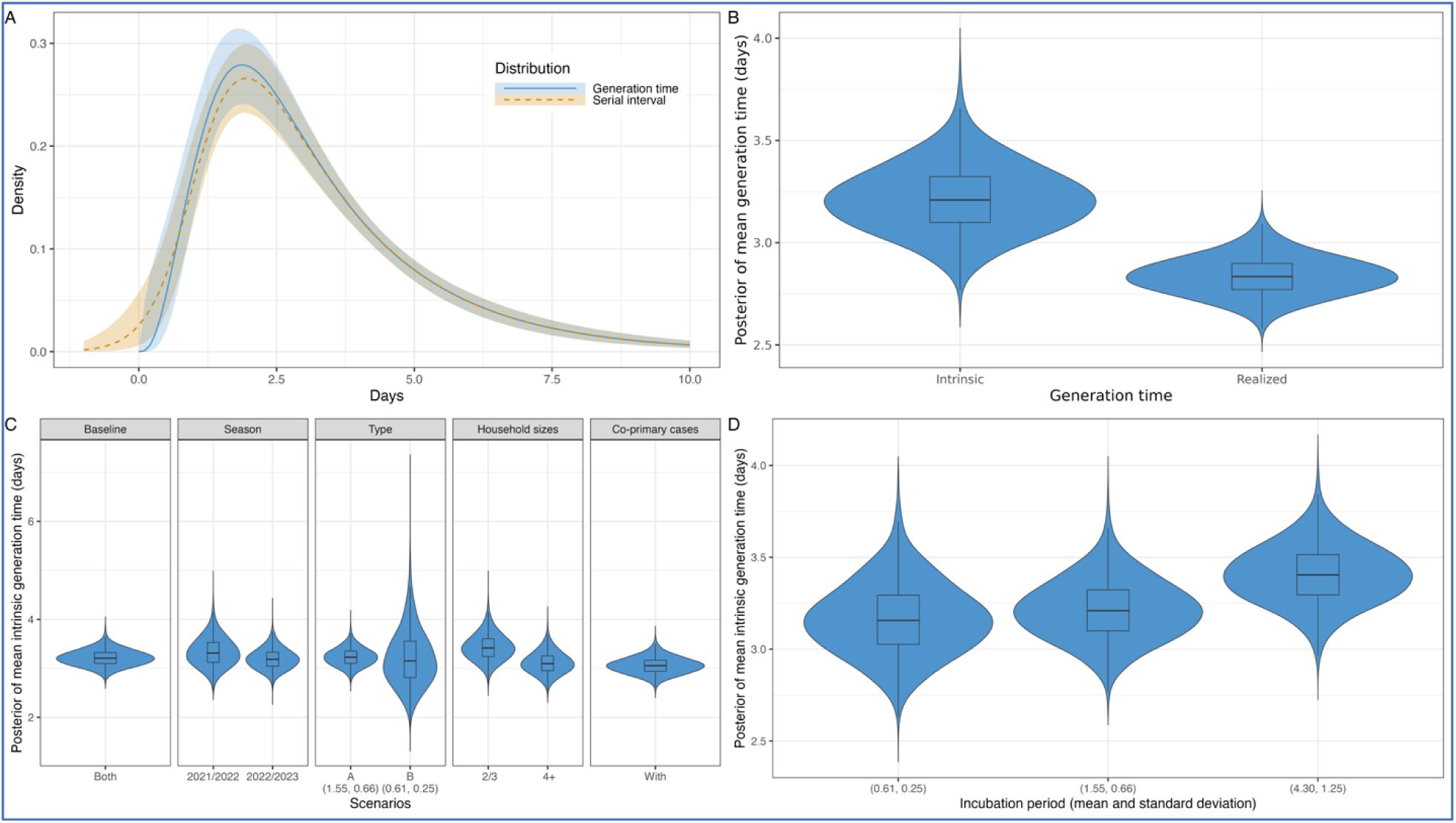
(A) Distributions of intrinsic generation time and serial interval using the posterior samples. The lines represent the median, and the shaded areas denote the 95% credible intervals (CrI). The blue color represents the intrinsic generation time distribution, while the orange color represents the serial interval distribution. (B) Posterior distribution of mean intrinsic and realized household generation time. (C) Posterior distributions of mean intrinsic generation time across seasons, virus types, household sizes, and with multiple co-primary cases. The incubation period, derived from influenza A, had a mean of 1.55 days and a standard deviation (SD) of 0.66 days (Lessler, et al. 2009). Only for influenza B, we assumed the shorter incubation period to yield a mean of 0.61 days and a standard deviation (SD) of 0.25 days (Lessler, et al. 2009). (D) Posterior distributions of mean intrinsic generation time estimated using the full dataset across different incubation periods.

**Table 2.** Posterior mean (95% CrIs) of estimates in primary analysis using the full dataset. The incubation period, derived from influenza A, had a mean of 1.55 days and a standard deviation (SD) of 0.66 days (Lessler, et al. 2009). The relative infectiousness of asymptomatic infected individuals compared with symptomatic infected individuals was assumed to be 0.57 (Tsang, et al. 2023).

|  | Mean | SD |
| --- | --- | --- |
| Intrinsic generation time (days) | 3.2 (2.9-3.6) | 2.1 (1.8-2.5) |
| Realized household generation time (days) | 2.8 (2.7-3.0) | 1.6 (1.5-1.8) |
| Serial interval (days) | 3.2 (2.8-3.5) | 2.2 (1.8-2.6) |

We found no substantial differences in the mean intrinsic generation time estimates across multiple data stratifications (Figure 1C and Supplemental Table S1). The overlapping indices for both 2021/2022 and 2022/2023 were high at 71% and 87%, respectively, aligning with the primary analysis above. Influenza A data showed a notable high overlapping index of 94%, reflecting its dominance, as influenza A was identified in 83% of the individuals. Conversely, using the data exclusively from influenza B yielded a similar mean but with a wider credible interval due to the smaller sample size, i.e., influenza B was identified in only 17% of the individuals, resulting in a lower overlapping index of 47%. Upon examining household sizes, although we found slightly longer mean intrinsic and realized household generation times in smaller households compared to larger ones, the overlapping index for household sizes of 2 or 3 members and 4 or more members were moderately high at 61% and 74%, respectively. Incorporating households with co-primary cases remained consistent with a moderately high overlapping index of 64%, indicating the similarity between exclusion and inclusion of multiple co-primary cases.

The mean intrinsic generation time exhibited limited sensitivity to variations in the incubation period (Figure 1D and Supplemental Table S2). In the primary analysis shown above, we used an incubation period with a mean of 1.55 days and a standard deviation (SD) of 0.66 days by fitting previously published estimates (Lessler, et al. 2009) to a gamma distribution (Supplemental Figure S1). When considering a shorter incubation period, which yielded a mean of 0.61 days and a SD of 0.25 days (Lessler, et al. 2009), the mean intrinsic generation time remained unchanged with an overlapping index of 86%. Conversely, with a longer incubation period, which yielded a mean of 4.30 days and a SD of 1.25 days (Tuite, et al. 2010), the mean intrinsic generation time increased slightly with an intermediate overlapping index of 56%.

Our estimates were not sensitive to changes in the relative infectiousness of asymptomatic infected individuals, due to the limited number of asymptomatic infected individuals in this study (Supplemental Figure S6). We found no substantial differences in the mean intrinsic generation time estimates (Supplemental Table S2), as indicated by overlapping indices of 94% and 97% when using the values of 0.11 and 1.54 compared to the primary value of 0.57 (Tsang, et al. 2023).

### Sensitivity of other transmission parameters to the incubation period

In our sensitivity analyses, where we varied the assumed incubation period from the mean of 1.55 days and SD of 0.66 days, we found significant influences on several crucial pre-symptomatic transmission parameters and the duration of various symptomatic infectious stages (Table 3 and Supplemental Figure S5).

**Table 3.** Posterior mean (95% CrIs) of estimates of generation time and transmission parameters given different assumed incubation periods. The primary incubation period, derived from influenza A, had a mean of 1.55 days and a standard deviation (SD) of 0.66 days (Lessler, et al. 2009). The shorter incubation period, derived from influenza B, yielded a mean of 0.61 days with a SD of 0.25 days (Lessler, et al. 2009), while the longer incubation period, derived from influenza A(H1N1)pdm09, had a mean of 4.30 days with a SD of 1.25 day (Tuite, et al. 2010). The relative infectiousness of asymptomatic infected individuals compared with symptomatic infected individuals was assumed to be 0.57 (Tsang, et al. 2023).

| Incubation period | Shorter | Primary | Longer |
| --- | --- | --- | --- |
| Mean intrinsic generation time (days) | 3.2 (2.8-3.6) | 3.2 (2.9-3.6) | 3.4 (3.1-3.7) |
| Proportion of transmission before symptomatic onset | 0.03 (0.00-0.06) | 0.16 (0.07-0.25) | 0.76 (0.65-0.87) |
| Ratio of pre-symptomatic and symptomatic transmission rates | 0.6 (0.1-1.7) | 0.7 (0.2-1.9) | 1.3 (0.5-3.2) |
| Latent period (days) | 0.4 (0.2-0.6) | 0.9 (0.2-1.4) | 0.9 (0.2-1.6) |
| Pre-symptomatic infectious period (days) | 0.2 (0.0-0.4) | 0.7 (0.2-1.3) | 3.4 (2.7-4.1) |
| Symptomatic infectious period (days) | 2.6 (2.3-3.0) | 2.0 (1.7-2.4) | 1.2 (0.7-1.9) |

Notably, given the shorter incubation period (mean of 0.61 days and SD of 0.25 days), the proportion of transmission before symptomatic onset was lower at 3% (95% CrI: 0-6%), and the ratio of pre-symptomatic to symptomatic transmission rates indicated a lower relative infectiousness of symptomatic infected individuals before symptom onset compared to after. This indicates that the majority of transmission occurred after individuals developed symptoms. Consequently, this was reflected in a shorter latent period of 0.4 (95% CrI: 0.2-0.6) days and pre-symptomatic infectious period of 0.2 (95% CrI: 0.0-0.4) days, or a longer symptomatic infectious period of 2.6 (95% CrI: 2.3-3.0) days.

Conversely, given the longer incubation period (mean of 4.30 days and SD of 1.25 days), the proportion of transmission before symptomatic onset was higher at 76% (95% CrI: 65-87%), and the ratio of pre-symptomatic to symptomatic transmission rates was higher. This resulted in a longer latent period of 0.9 (95% CrI: 0.2-1.6) days and pre-symptomatic infectious period of 3.4 (95% CrI: 2.7-4.1) days, while the symptomatic infectious period was shorter at 1.2 (95% CrI: 0.7-1.9) days.

## Discussion

### Estimates of generation time

This study employed a Bayesian data augmentation approach (Hart, Maini and Thompson 2021, Hart, Abbott, et al. 2022, Hart, Miller, et al. 2022) to estimate both intrinsic and realized generation times using data collected from a U.S. household study during the post COVID-19 pandemic influenza seasons, 2021/2022 and 2022/2023. Our findings indicate that the intrinsic generation time, reflecting transmission dynamics within community settings, ranged from 2.9 to 3.6 days, while the realized household generation time, restricted to household settings, ranged from 2.7 to 3.0 days. These estimates of the generation time for influenza fall within the uncertainty bounds of pre-pandemic studies, including directly using viral shedding data (Carrat, et al. 2008) and other contact tracing data (Fraser, et al. 2009) (te Beest, et al. 2013) (Lau, et al. 2015), with estimates varying between 2 and 4 days, suggesting that there has not been substantial change since the 2009 H1N1 influenza pandemic. Additionally, the overlapping indices higher than 70% suggested no substantial differences between the two influenza seasons.

Both seasons of this study were atypical, being the first seasons since the COVID-19 pandemic, during which the immunity to influenza had potentially decreased. The 2022/2023 season, in particular, experienced an early influenza activity peak along with RSV and COVID-19 outbreaks. Despite these unusual circumstances, both seasons dominated by influenza A(H3N2) were tested in our sensitivity analyses, which were based on different parameter assumptions. The generation time estimates remained similar to those from earlier studies, suggesting that virus transmission dynamics within households have not changed substantially and may not vary widely between types. However, further work is needed to fully explore estimates for influenza that were less prevalent in this study (i.e., influenza B and A(H1N1)).

Our finding that the realized household generation time was shorter than the intrinsic generation time could be attributed to the depletion of susceptible individuals over time. As household members become infected and develop immunity, although individuals may still be infectious, there are no susceptible contacts still exposed to each case. This depletion terminates transmission chains, diminishing the potential for further infections. This process, along with factors such as closer proximity and longer exposure times inherent to household settings, can increase the chance of transmission within households, leading to a shorter observed generation time.

Our updated estimates, particularly for the intrinsic generation time, may be useful for ongoing modeling efforts which require estimated generation times, such as real-time influenza Rt estimation (Centers for Disease Control and Prevention 2024) (Centers for Disease Control and Prevention 2024) (Gostic, et al. 2020). Our estimates are slightly shorter than the serial interval estimated by Cowling et al. (Cowling, et al. 2009) at 3.6 days (95% confidence interval, CI: 2.9-4.3). This shorter interval suggests a more rapid spread, potentially leading to higher estimated Rt and emphasizes the need for prompt and effective interventions to control transmission.

### Reliability of other transmission parameters

Comparing our other parameter estimates with prior research, we found the mean serial interval to be 3.2 days, within the 95% CI of 2.9 to 4.3 days reported by Cowling et al. (Cowling, et al. 2009). Our estimate also falls within the 3-to-4-day range of uncertainty reported in previous household studies (Cauchemez, Donnelly, et al. 2009, Petrie, et al. 2013, Levy, et al. 2013, Xu, et al. 2015, Cowling, et al. 2010, Suess, et al. 2012, Boëlle, et al. 2011, Tsang, et al. 2016).

We estimated a latent period of less than a day, which is shorter than the 1 to 3 days reported in other studies (Tuite, et al. 2010) (Cori, et al. 2012). It is possible that this shorter latent period could be influenced by undocumented exposures outside households. For instance, both the index case and infected household member may have been exposed to influenza elsewhere, with the index case developing symptoms before the household member. Consequently, when the household member becomes sick, we attribute it to the index case within the household, but this infection could have originated from previous exposure outside the household, which has not been accounted for in this analysis. Nonetheless, we excluded households with multiple co-primary cases to reduce these effects, ensuring accurate assessment of transmission dynamics within each household.

Furthermore, there was substantial uncertainty in our estimates for the symptomatic infectious period, which ranged from 1 to 3 days, given different assumed incubation periods. Likewise, there has been a wide range of estimates in earlier studies, including those less than a day (Cori, et al. 2012) and more than 3 days (Tuite, et al. 2010) (Cauchemez, Carrat, et al. 2004).

The estimates of the pre-symptomatic transmission parameters and the duration of various symptomatic stages should be interpreted with caution due to the inherent uncertainty in accurately estimating the incubation period. Our estimates incorporated values from various studies based on different types or subtypes of influenza (Lessler, et al. 2009) (Tuite, et al. 2010). This variability in the incubation period contributes to a wide range of pre-symptomatic transmission parameters. Without a reliable input for the incubation period, accurately determining these transmission dynamics becomes challenging.

### Implications for preventing transmission

Understanding the proportion of transmission that occurs prior to symptoms is critical to informing effective disease control strategies and assessing the potential impact of post-symptomatic mitigation measures, such as isolation of cases. We estimated that between 3% and 76% of transmission may occur before a person develops symptoms. This wide range was influenced by our assumptions of the incubation period, which were taken from a variety of previously published estimates. Longer incubation period assumptions yielded higher estimates of the percentage of transmission that occurred before symptoms. Similarly, attempts to estimate individual-level pre-symptomatic transmission using viral kinetics data have revealed substantial heterogeneity (Morris, et al. 2024). This highlights that pre-symptomatic transmission of influenza does occur, aligning with findings for other respiratory pathogens like SARS-CoV-2 (Buitrago-Garcia, et al. 2022).

Given the wide range of pre-symptomatic transmission, relying solely on isolation of symptomatic individuals may reduce but not eliminate influenza transmission. Although isolation measures initiated after symptom onset are likely to mitigate at least some influenza spread, given the relatively low levels of asymptomatic infection and pre-symptomatic transmission among the majority of individuals, there remains large heterogeneity (Morris, et al. 2024). A layered approach, including isolation of ill or infected people, maintaining good respiratory hygiene, and promoting influenza vaccination, may be most effective to reduce transmission within households (Centers for Disease Control and Prevention 2024). This underscores the importance of vaccination as the primary recommendation to prevent influenza-associated morbidity and mortality, especially for individuals at increased risk of influenza complications.

### Modeling details and limitations

While our reliance on household data might introduce limitations, such as the presence of multiple co-primary cases, sensitivity analyses confirmed the robustness of our generation time estimates to various data stratifications and model assumptions. Nevertheless, there are several limitations to our study.

First, we did not account for vaccination status. This omission is less likely to impact our estimates for influenza given similarities in viral load dynamics between infected vaccinated and unvaccinated individuals (Morris, et al. 2024) (Suess, et al. 2012). Nevertheless, it is possible that different vaccination statuses could be associated with different generation time estimates.

Second, we did not account for potential exposures outside households. When we excluded household members who did not have at least two valid PCR tests, the household sizes were reduced in our analysis. While this might not directly affect our generation time estimates, it could lead to an overestimation of overall infectiousness due to the absence of unobserved uninfected members.

Third, it is essential to acknowledge the influence of the COVID-19 pandemic on our influenza data. Generalizing these findings to pre-pandemic or post-pandemic periods should be done with caution. During the period when this study was conducted, individuals may have been more likely to adopt preventive measures against transmission within the home, such as self-isolating, practicing good respiratory and hand hygiene, wearing masks, and reducing contact with household members. These non-pharmaceutical interventions, alongside changes in human behavior and heightened awareness of infection control, could have impacted the spread of influenza.

## Conclusions

Through comprehensive data collected during the 2021/2022 and 2022/2023 influenza seasons in the U.S., we provide updated estimates of the generation time, essential for informing influenza modeling and public health strategies. Despite the significant changes in public behavior and preventive measures due to the COVID-19 pandemic, our study did not detect substantial changes in the generation time of influenza since the 2009 influenza pandemic. This finding is particularly significant given that the study period followed the extreme measures implemented to prevent COVID-19, which also reduced the transmission of influenza and other respiratory infections. Our findings contribute to our understanding of influenza transmission dynamics within households and underscore the importance of ongoing research for effective outbreak management.

## Data Availability

The household data are available upon reasonable request and upon completion of required approvals. The R code for estimating the generation time is available at https://github.com/CDCgov/influenza-generation_time-us.

## Author contributions

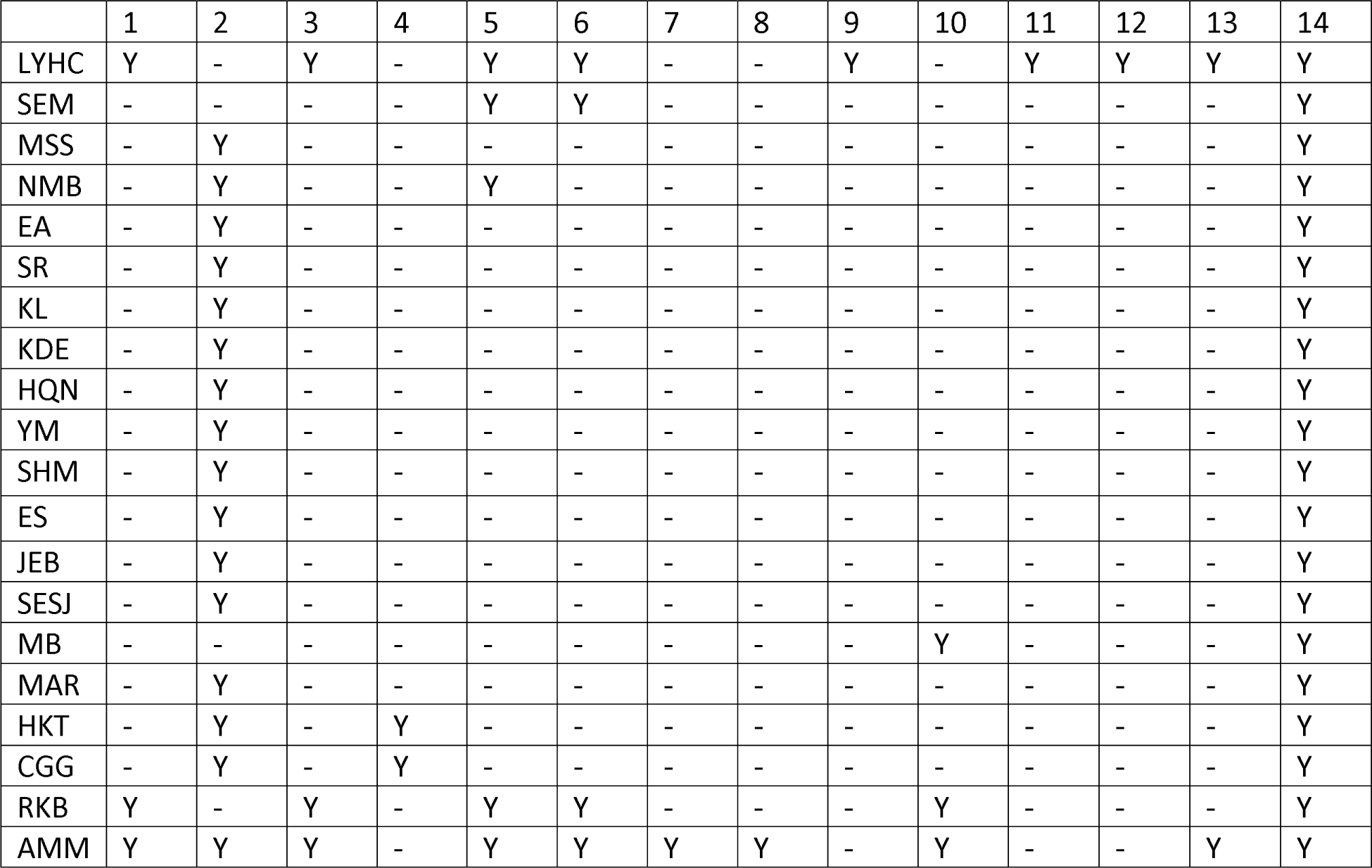

… choose from the following items: https://credit.niso.org / https://www.elsevier.com/researcher/author/policies-and-guidelines/credit-author-statement

1. Conceptualization
2. Data curation
3. Formal analysis
4. Funding acquisition
5. Investigation
6. Methodology
7. Project administration
8. Resources
9. Software
10. Supervision
11. Validation
12. Visualization
13. Roles/Writing - original draft
14. Writing - review & editing.

## Acknowledgments

- The authors thank the following members of the Respiratory Virus Transmission Network – Sentinel (RVTN-S) study teams:

a. Vanderbilt University Medical Center: Chris Lindsell, Judy King, John Meghreblian, Samuel Massion, Brittany Creasman, Lauren Milner, Andrea Stafford Hintz, Jorge Celedonio, Ryan Dalforno, Maria Catalina Padilla-Azain, Daniel Chandler, Paige Yates, Brianna Schibley-Laird, Alexis Perry, Ruby Swaimn, Mason Speirs, Erica Anderson, Suryakala Sarilla, Amelia Dodds, Dayton Marchlewski, Timothy Williams, Afan Swan, Onika Abrams, Jackson Resser, Ine Sohn, Cara Lwin, Hsi-nien (Jubilee) Tan, Stephen Yeargin, James Grindstaff, Heather Prigmore, Jessica Lai, Zhouwen Liu, James D. Chappell, Marcia Blair, Rendie E. McHenry, Bryan P. M. Peterson, Lauren J. Ezzell.
b. Columbia University: Lisa Saiman, Raul A. Silverio Francisco, Anny L. Diaz Perez, Ana M. Valdez de Romero.
c. Stanford University: Rosita Thiessen, Marcela Lopez, Alondra A. Aguilar, Emma Stainton, Grace K-Y. Tam, Jonathan Altamirano, Leanne X. Chun, Rasika Behl, Samantha A. Ferguson, Yuan J. Carrington, Frank S. Zhou.
d. Marshfield Clinic Research Institute: Edward A. Belongia, Hannah Berger, Vicki Moon, Gina Burbey, Leila Deering, Brianna Freund, Garrett Heuer, Sarah Kopitzke, Carrie Marcis, Jennifer Meece, Jennifer Moran, DeeAnn Hertel, Joshua Petrie, Miriah Rotar, Carla Rottscheit, Elisha Stefanski, Sandy Strey, Melissa Strupp.
e. University of Arizona: Ferris Alaa Ramadan, Flavia Maria Nakayima Miiro, Josue Ortiz, Mokenge Ndiva Mongoh.
f. University of North Carolina: Ayla Bullock, Amy Yang, Quenla Haehnel, Jessica Lin, Julienne Reynolds, Katherine “Katie” Murray, Miriana Moreno Zivanovich, Anna McShea, Brittney Figueroa, Melody Liu.
g. University of Colorado: Kathleen Grice, Cameron Bendalin, Sonia Chavez, Jolie Granger.
- We also acknowledge the households for their participation.
- L. Y. H. C. thanks the CDC Steven M. Teutsch Prevention Effectiveness (PE) Fellowship.

## Disclaimer

The conclusions, findings, and opinions expressed by authors contributing to this article do not necessarily reflect the official position of the U.S. Department of Health and Human Services, the Public Health Service, the Centers for Disease Control and Prevention, or the authors’ affiliated institutions.

## Declaration of Generative AI and AI-assisted technologies in the writing process

During the preparation of this work the authors used ChatGPT in order to enhance the clarity, coherence, and correctness of the writing, and to check for grammatical errors. After using this tool, the authors reviewed and edited the content as needed and take full responsibility for the content of the publication.

## Supplementary material

### The incubation period distribution

The incubation period distribution was modeled using estimates for influenza A from a systematic review by Lessler et al. (Lessler, et al. 2009), with a mean of 1.55 days and a standard deviation (SD) of 0.66 days. These estimates were fitted to a gamma distribution to characterize the distribution of the incubation period (Supplemental Figure S1).

**Figure S1.**
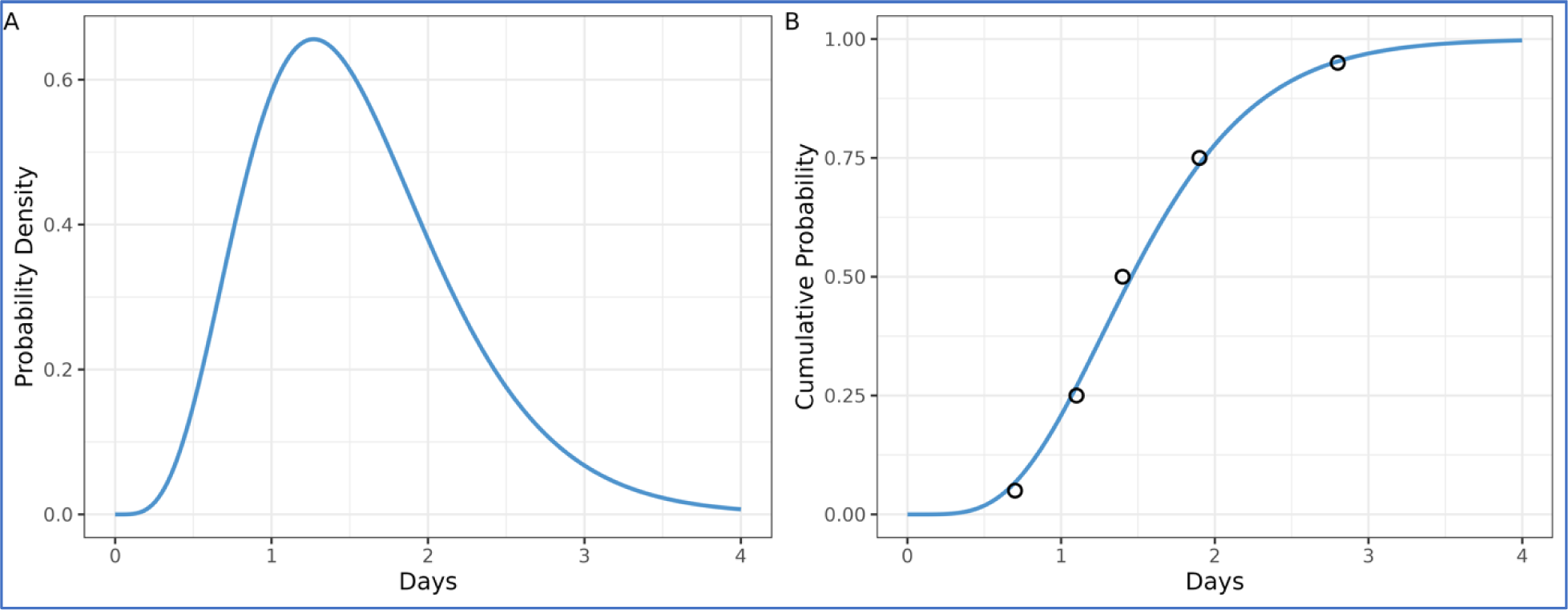
Incubation period distribution. The black circles and blue lines represent the data (Lessler, et al. 2009), and the (A) cumulative distribution function and (B) probability density function of a gamma distribution fitted to the data.

### The observed household serial interval of single infection pairs

We found that the observed household serial interval, calculated without modeling, solely using data from households with single infection pairs (i.e., single primary case to single secondary case) and without potential transmission chains, had a mean of 3.7 days (and a SD of 2.3 days). This was longer than the mean intrinsic serial interval of 3.2 (95% CrI: 2.8-3.5) days when considering households of all sizes with all potential transmission chains (Table 2). This does not necessarily indicate that the intrinsic value was shorter than the realized household one. Rather, it is mainly due to the restriction of single infection pairs or mostly smaller household sizes of 2 members.

In the main text, we found slightly longer mean intrinsic and realized household generation times in smaller households compared to larger ones (Figure 1C and Supplemental Table S1). Larger households with more exposure and potential transmission chains could have a shorter interval, while smaller households could have a longer interval.

### Specification of parameters for the mechanistic model

In the mechanistic model (Hart, Abbott, et al. 2022), two parameters, namely the ratio of the mean latent and incubation period and the mean symptomatic infectious period, were estimated directly (Supplemental Figure S2), while the proportion of transmission before symptomatic onset was calculated by weighting the pre-symptomatic period by the ratio of pre-symptomatic and symptomatic transmission rates and dividing it by the sum of the (pre-symptomatic and symptomatic) infectious periods. The mean latent and mean pre-symptomatic periods were calculated by dividing the incubation period by the ratio of mean latent and incubation period.

**Figure S2.**
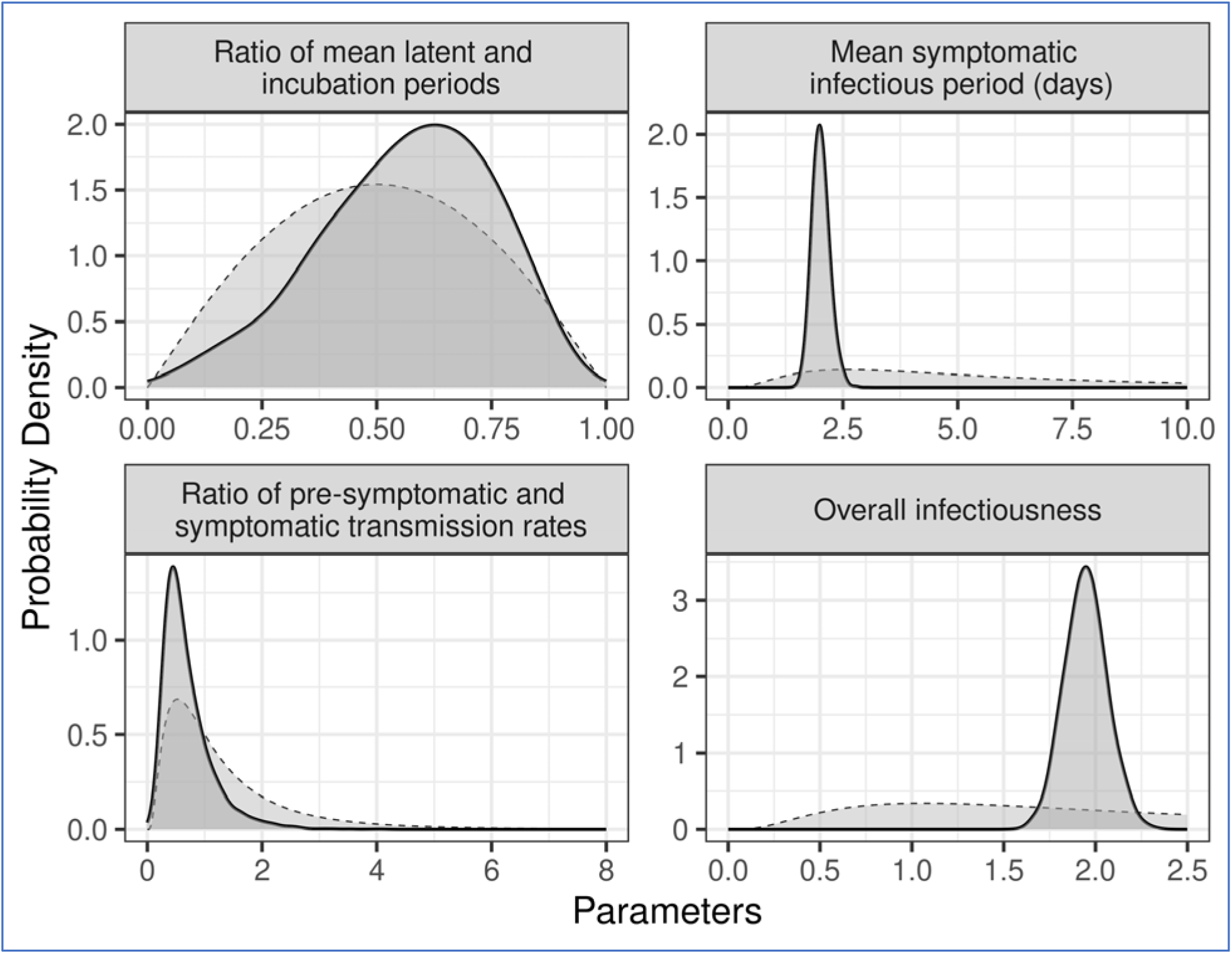
Posterior and prior distributions of estimated parameters. Solid and dashed lines represent posterior and prior distributions, respectively.

### Variability in estimates across data stratifications

Although the generation time or serial interval of influenza B may be longer than that of influenza A (Levy, et al. 2013), this was not the case in our findings from the two seasons (Supplemental Table S1, Figure S3 and S4). However, we note that the mean intrinsic generation time exhibited a wider credible interval when using data exclusively from influenza B compared to influenza A, which likely reflects the dominance of influenza A during the study timeframe and the smaller sample size of influenza B.

**Table S1.** The posterior mean (95% CrIs) of mean intrinsic generation time across seasons, virus types, household sizes, and with multiple co-primary cases. The incubation period, derived from influenza A, had a mean of 1.55 days and a standard deviation (SD) of 0.66 days (Lessler, et al. 2009). Only for influenza B, we assumed the shorter incubation period to yield a mean of 0.61 days and a standard deviation (SD) of 0.25 days.

| Data stratifications | Mean intrinsic generation time (95% CrIs) | Overlapping index (% compared to the primary analysis) |
| --- | --- | --- |
| All data excluding households with multiple co-primary cases (primary analysis in Table 1) | 3.2 (2.9-3.6) | 100 |
| Season 2021/2022 | 3.3 (2.8-4.0) | 71 |
| Season 2022/2023 | 3.2 (2.8-3.6) | 87 |
| Influenza A | 3.2 (2.9-3.6) | 94 |
| Influenza B | 3.2 (2.3-4.5) | 47 |
| Household size of 2 or 3 | 3.4 (2.9-4.0) | 61 |
| Household size of 4 or greater | 3.1 (2.7-3.6) | 74 |
| All data including households with multiple co-primary cases | 3.1 (2.7-3.4) | 64 |

**Figure S3.**
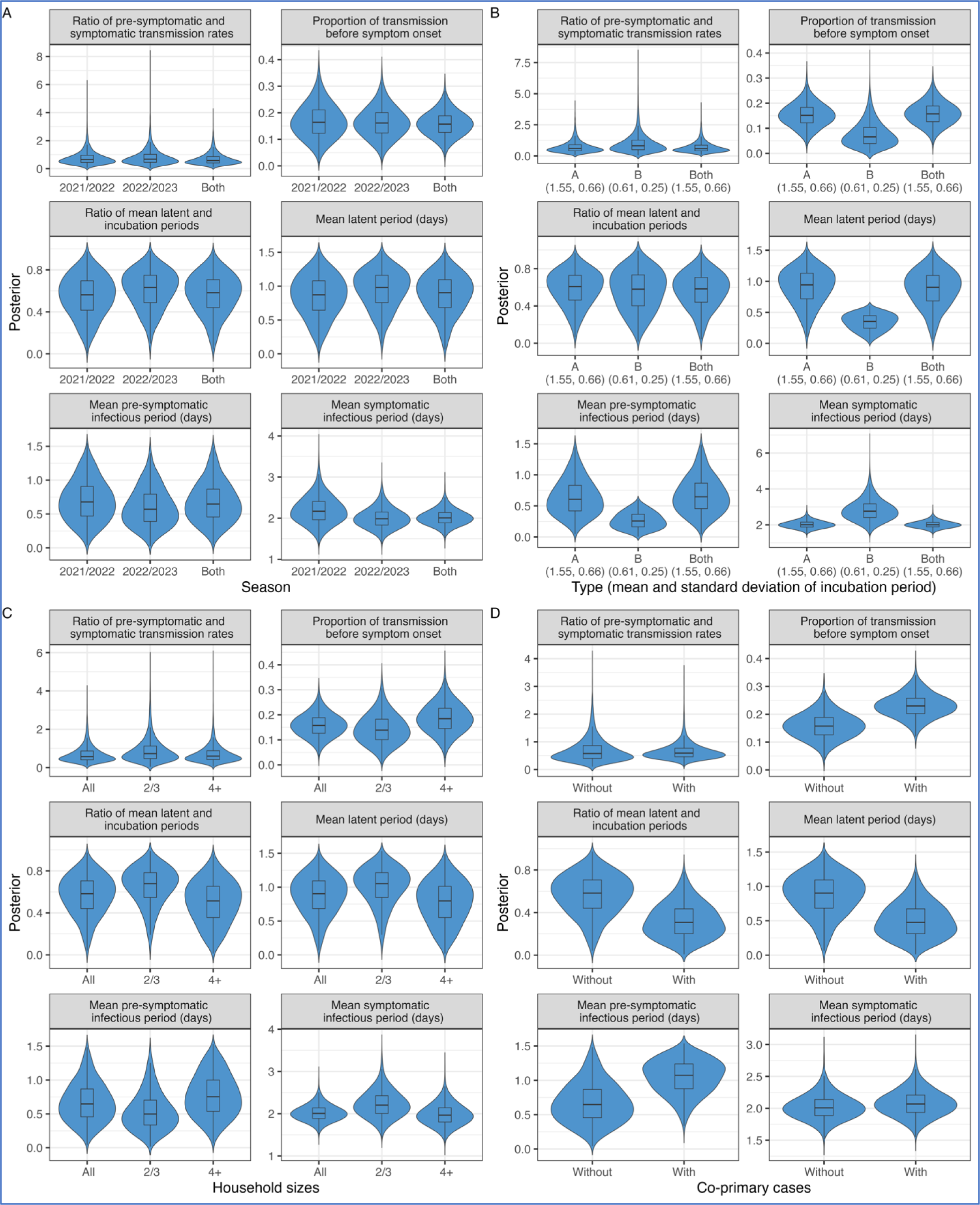
Posterior distributions of parameters across data stratifications: (A) seasons, (B) virus types, (C) household sizes, and (D) with multiple co-primary cases.

**Figure S4.**
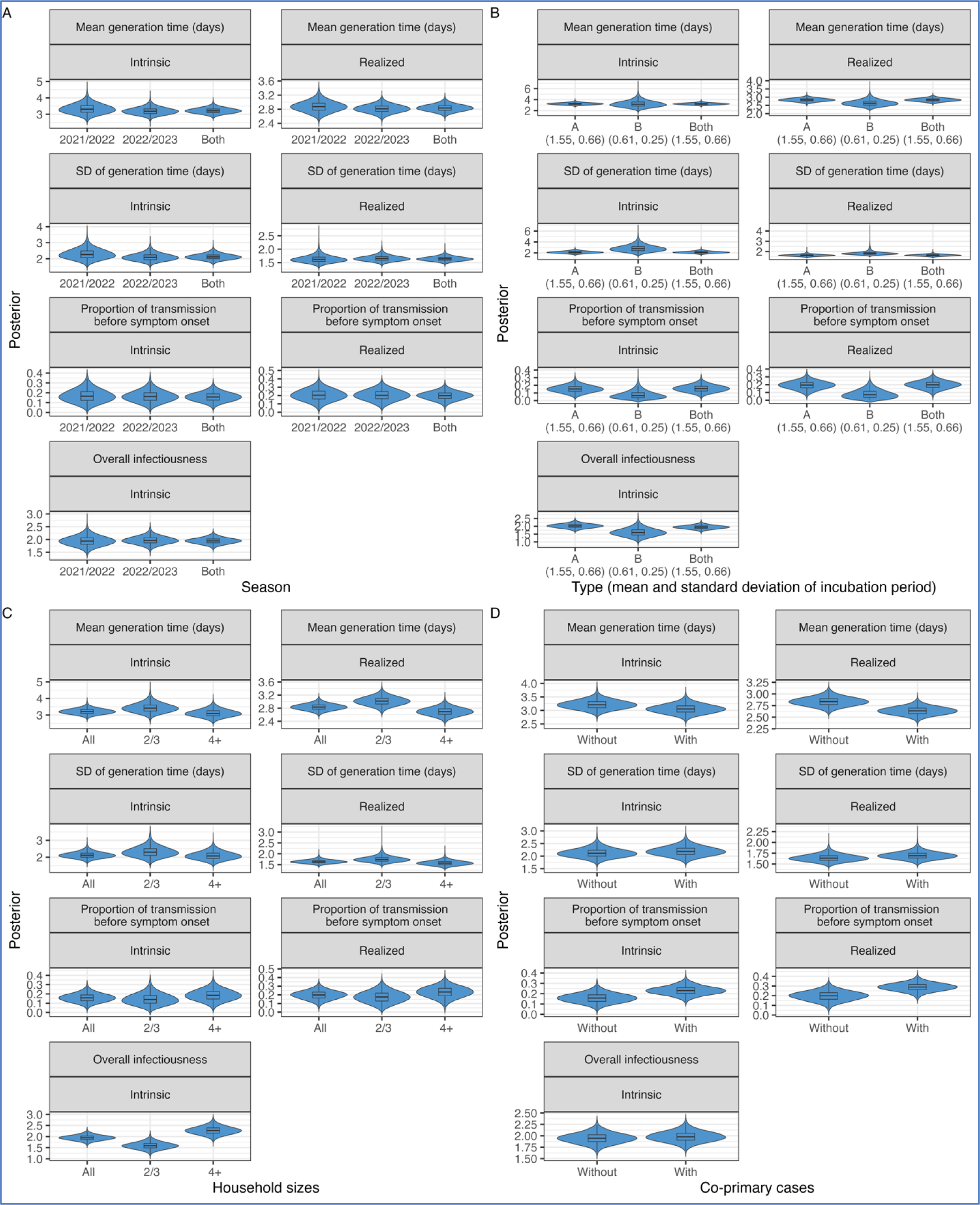
Posterior distributions of parameters across data stratifications: (A) seasons, (B) virus types, (C) household sizes, and (D) with multiple co-primary cases.

### Sensitivity analyses

Similar to the sensitivity analyses using the full dataset, we found that the incubation period had a limited effect on the intrinsic generation time when exclusively using data from households circulating influenza A (Supplemental Figure S6, Panel B) or households circulating influenza B (Supplemental Figure S6, Panel C).

Consistent with the previous study (Hart, Abbott, et al. 2022), assuming a higher relative infectiousness of asymptomatic infected individuals resulted in slightly lower estimates of the overall infectiousness of infectors (Supplemental Figure S6, Panel D).

**Table S2.** The posterior mean (95% CrIs) of mean intrinsic generation time given different incubation periods or relative infectiousness of asymptomatic infected individuals. The primary incubation period, derived from influenza A, had a mean of 1.55 days and a standard deviation (SD) of 0.66 days (Lessler, et al. 2009). For the shorter incubation period derived from influenza B, we assumed a mean of 0.61 days and a SD of 0.25 days (Lessler, et al. 2009). For the longer incubation period derived from influenza A(H1N1)pdm09, we assumed a mean of 0.61 days and a SD of 0.25 days (Tuite, et al. 2010).

| Sensitivity analyses | Mean intrinsic generation time (95% CrIs) | Overlapping index (% compared to the primary analysis) |
| --- | --- | --- |
| Primary analysis (in Table 1) | 3.2 (2.9-3.6) | 100 |
| Longer incubation period | 3.2 (2.8-3.6) | 86 |
| Shorter incubation period | 3.4 (3.1-3.7) | 56 |
| Lower relative infectiousness | 3.2 (2.9-3.6) | 94 |
| Higher relative infectiousness | 3.2 (2.9-3.6) | 97 |

**Figure S5.**
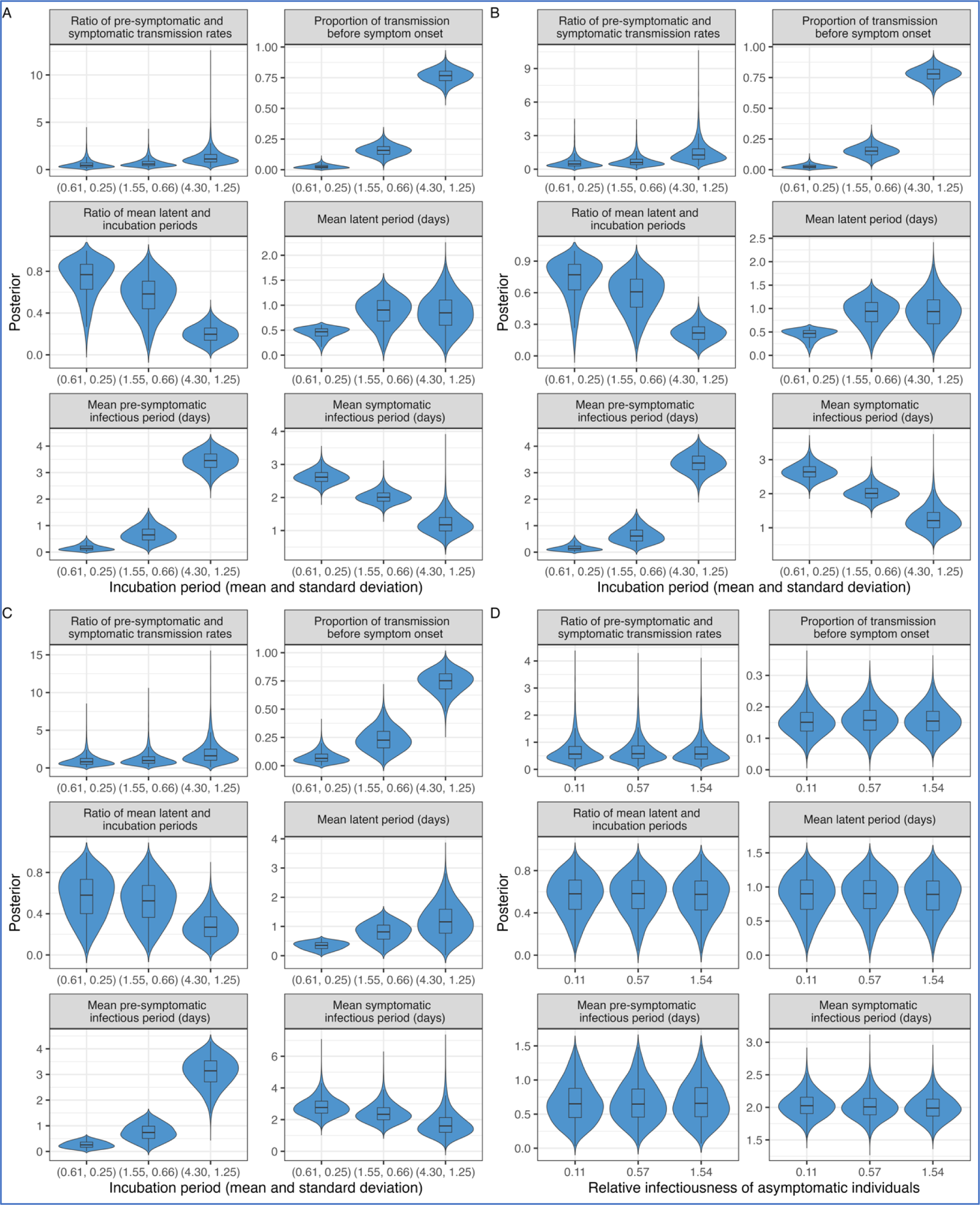
Posterior distributions of parameters given different assumptions: (A-C) incubation periods, and (D) relative infectiousness of asymptomatic infected individuals. Panel (A) presents results obtained using data from households with both influenza A and B, whereas Panels (B) and (C) present results obtained using data solely from households with influenza A and B, respectively.

**Figure S6.**
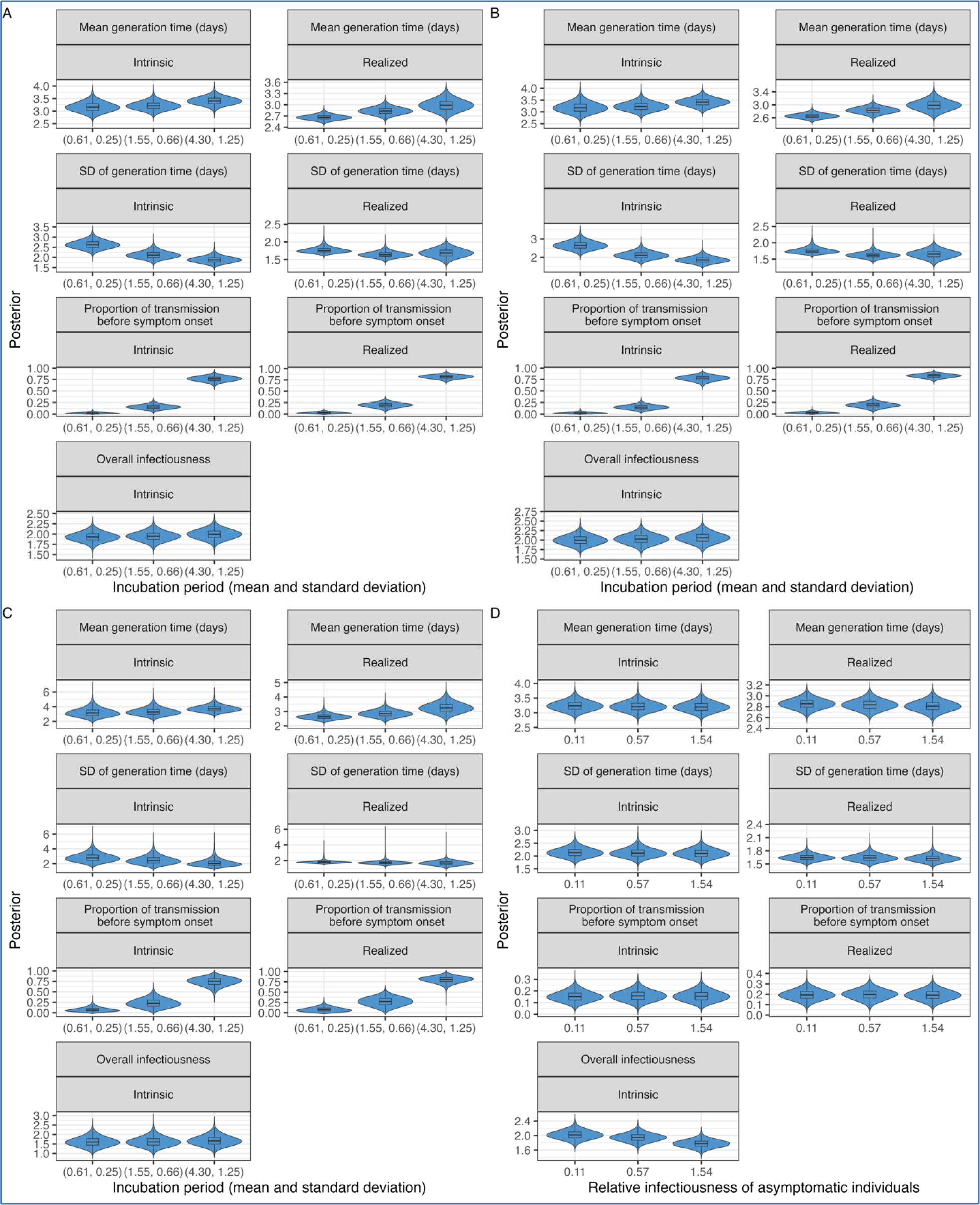
Posterior distributions of parameters given different assumptions: (A-C) incubation periods, and (D) relative infectiousness of asymptomatic infected individuals. Panel (A) presents results obtained using data from households with both influenza A and B, whereas Panels (B) and (C) present results obtained using data solely from households with influenza A and B, respectively.

